# Forecasting Minute-by-Minute Stress, Anxiety, and Affective States Using Time-Series Analysis of Wearable Sensor Data

**DOI:** 10.64898/2026.04.29.26352047

**Authors:** Destiny Pounds, Vibhhuti Gupta, Himanshu Tripathi, Subash Neupane

**Affiliations:** Department of Computer Science & Data Science, Meharry Medical College, Nashville, USA; Department of Biostatistics & Data Science, University of Texas Medical Branch, Galveston, USA; Department of Computer Science, University of Alabama, Tuscaloosa, Country

**Keywords:** Wearable sensors, stress detection, anxiety prediction, affective computing, time-series, HRV, EDA, machine learning

## Abstract

This paper focuses on forecasting minute-by-minute stress, anxiety, and affective states using wearable sensor data. It addresses mental health as a growing concern and the limitations of traditional assessment methods. A time-series machine learning framework was developed using electrodermal activity (EDA) and heart rate variability (HRV) features from the WESAD dataset. Models were trained and evaluated for minute-by-minute prediction of self-reported psychological states. Both classification (stress, anxiety) and regression models (affect) were explored comparing time-series and static approaches. Findings support the feasibility of real-time, personalized mental health monitoring using wearable devices and their potential for timely interventions in clinical or remote settings. The paper demonstrates how temporal modeling can enhance emotional state prediction and inform future research and system development.

## I. Introduction

Mental health has become a growing public health concern, affecting millions of adults in the United States each year. According to the 2022 SAMHSA Annual Report, 59.3 million people (23.1% of the US population) live with mental illness, with the highest rates among young adults aged 18–25 [1]. In 2022, 18.2% of adults reported anxiety symptoms and 21.4% reported depressive symptoms, with the highest rates among young adults, women, rural populations and individuals of lower economic status [2]. Chronic stress is estimated to be a major risk factor for 75–90% of all diseases [3], influencing not only psychological health but also physical conditions such as immune suppression, cardiovascular disease, and neurodegeneration [4].

Stress is particularly relevant to anxiety disorders, which often occur concurrently with other disorders [5]. Despite its significant influence, stress is rarely monitored in real time, especially in relation to its impact on mental health. Traditional diagnostic approaches rely heavily on clinical interviews, behavioral observations, and standardized self-report surveys [6]. These tools are limited by their retrospective nature, risk of subjective bias, and inconvenience. These limitations often result in delayed diagnosis and missed opportunities for early intervention. Wearable devices containing biosensors can measure physiological signals such as electrodermal activity (EDA) and heart rate variability (HRV) in a continuous and non-invasive approach [7]. EDA and HRV signals are directly influenced by nervous system response and have been linked to emotional and cognitive states [8]. However, current models rarely leverage time-specific patterns in physiological data to predict self-reported psychological states [9]. This paper explores the potential use of wearable sensor technology in the realm of mental health care. By integrating subjective assessments with wearable sensor data, this work seeks to enhance stress detection models, making them more relevant for clinical applications. The primary goal is to design and evaluate machine learning models capable of forecasting an individual’s self-reported emotional state, specifically stress, anxiety, and affect in the upcoming minute, using physiological sensor data as input. Two modeling approaches are employed: (1) static models that predict emotional states from single one-minute data snapshots, and (2) time-series models that use sliding ten-minute windows to forecast the next minute’s state.

We hypothesize that time-series models utilizing aggregated EDA and HRV features from wearable sensors can forecast an individual’s stress levels, anxiety levels, and affective states for the upcoming minute more effectively than static prediction models. Based on this hypothesis, the following research questions guide this study:

**RQ1:** Can physiological signals from wearable devices be used to reliably forecast self-reported psychological states?

**RQ2:** Do time-series models outperform static models in predicting emotional states across participants? **RQ3:** Which physiological features are most predictive of self-reported stress, anxiety, and affect?

The remainder of this paper is organized as follows. Section II provides a brief literature review. Section III describes the dataset, data preprocessing pipeline, feature extraction methods, modeling approaches, and evaluation plan. Section IV presents the performance results of static and time-series models, provides visualizations, and interprets key findings including feature importance analysis and limitations. Finally, Section V summarizes the outcomes and offers recommendations for future research.

## II. Related Works

Previous studies have validated HRV and EDA as physiological indicators of stress with models achieving 70–90% accuracy in labs [10] but degrading to 60–80% in everyday scenarios [9]. A primary gap is limited generalizability due to lower signal quality and need for personalization [9]. Another gap is the limited integration of EDA with self-reported stress assessments [9]. Additionally, there has not been much focus on integrating stress and emotion detection systems [11]. This paper addresses these gaps by developing models that predict stress, anxiety, and affective states using both HRV and EDA integrated with self-reports from the WESAD dataset.

## III. Design and Methodology

### A. Dataset Overview

This study uses the WESAD dataset [11], which contains physiological data collected through wearable chest and wrist devices in a laboratory environment. The dataset includes data from 15 participants (12 male, 3 female) aged 24–35 who underwent four experimental conditions: Baseline (20-minute neutral recording), Amusement (humorous video clips, 392 seconds), Stress (modified Trier Social Stress Test with public speaking and mental arithmetic), and Meditation (seven-minute guided breathing). Self-reports were collected after each condition using PANAS, STAI, SAM, and SSSQ surveys. Figure 1 illustrates the overall workflow for this project, from raw signal ingestion through preprocessing, feature extraction, label mapping, modeling, and evaluation.

**Fig. 1:**
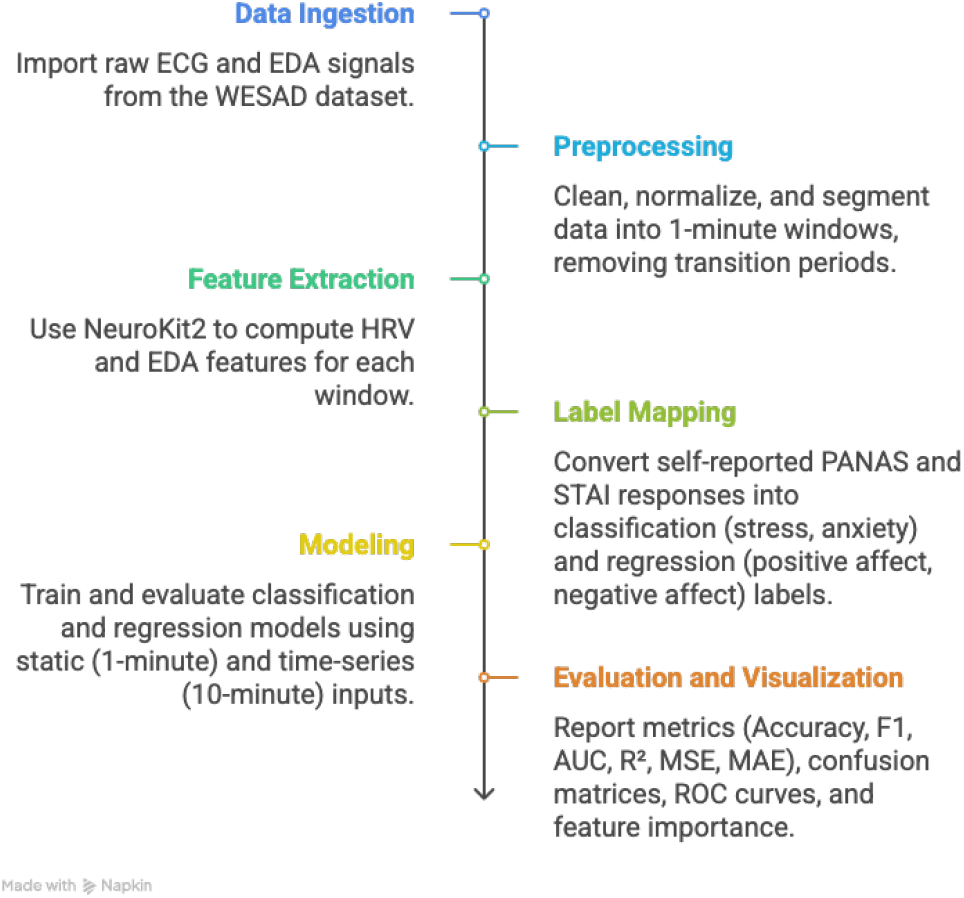
paper Workflow - shows data ingestion, preprocessing, feature extraction, label mapping, modeling, and evaluation pipeline

### B. Proposed Method

Two modeling approaches were implemented. *Static Models* treat each minute of physiological data as an independent sample and predict the corresponding self-report label with no temporal context. *Time-Series Models* use a sliding window of ten minutes of past physiological data and past labels to predict the upcoming minute’s label. Each minute of data was aggregated from raw EDA and ECG signals using NeuroKit2 [12]. Transition periods between conditions were excluded. The prediction tasks were: Stress Prediction (3-class classification from PANAS scores: 0 = No Stress, 1 = Low Stress, 2 = High Stress), Anxiety Prediction (3-class classification from modified 6-item STAI scores: 0 = No/Low, 1 = Moderate, 2 = High Anxiety), and Affective State Prediction (regression predicting continuous PA and NA scores from PANAS, range 10–50). For each task, multiple models were evaluated: Decision Tree, Logistic Regression (classification only), Random Forest, XGBoost, LightGBM, AdaBoost, SVM, and Linear Regression (regression only). Hyperparameter tuning was performed using GridSearchCV. Models were cross-validated using GroupKFold with *k* = 5 to ensure that all data from a given participant appeared in only one fold.

Figure 2 compares the two modeling strategies. The static model uses a single-minute snapshot of physiological features to predict the emotional state for that same minute. The time-series model uses a sliding window of the past ten minutes, including both physiological features and self-reported labels, to forecast the state in the next subsequent minute. The latter design more closely reflects real-world forecasting scenarios and leverages temporal patterns for improved prediction accuracy.

**Fig. 2:**
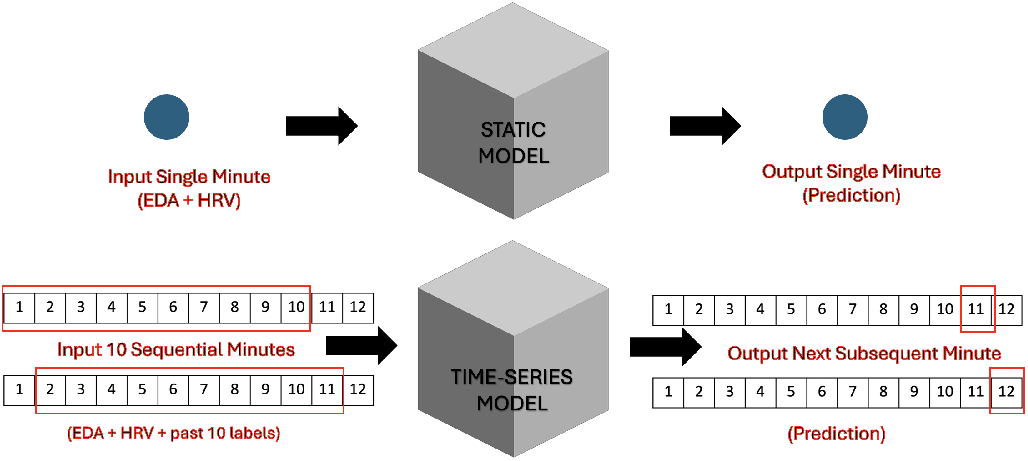
Modeling Approaches - compares static (single-minute input to single-minute output) vs. time-series (10-minute sliding window input to next-minute output) strategies

### C. Feature Extraction and Label Assignment

Physiological signals were collected using the RespiBAN chest device at 700 Hz. Only ECG and EDA channels were used. Raw signals were segmented into one-minute windows, cleaned, and normalized. Features were extracted using NeuroKit2 as listed in Table I, including:

**TABLE I:** Extracted Feature Set from Biosignals.

| Feature | Src | Description | Unit |
| --- | --- | --- | --- |
| SCL_Mean | EDA | Mean skin conductance | $\mu S$ |
| SCL_STD | EDA | Std. dev. of skin conductance | $\mu S$ |
| SCR_Peaks_Count | EDA | Skin conductance response count | Count |
| HRV_RMSSD | ECG | RMSSD of RR intervals | ms |
| HRV_SDNN | ECG | Std. dev. of RR intervals | ms |
| HRV_pNN50 | ECG | % RR intervals >50ms apart | % |
| HRV_MeanRR | ECG | Mean RR interval | ms |
| HRV_MedianRR | ECG | Median RR interval | ms |
| HRV_HF | ECG | High-freq. HRV power | $ms^2$ |
| HRV_LF | ECG | Low-freq. HRV power | ms |
| HRV_LF_HF_Ratio | ECG | LF/HF power ratio | — |
| HRV_SampEN | ECG | Sample entropy of RR intervals | — |

Stress labels were derived from PANAS (mapped: 1 → 0, 2–3 → 1, 4–5 → 2). Anxiety labels were based on 6-item STAI scores (4–9 → 0, 10–12 → 1, 13–24 → 2) [13], [14]. PA and NA scores were treated as continuous targets (range 10–50) [13]. Labels were validated by cross-referencing with SAM valence and arousal scores [15] as shown in Figure 3. Stress, anxiety, and NA labels were validated; however, PANAS PA scores did not reliably align with SAM valence, so PA prediction was excluded from final evaluation.

**Fig. 3:**
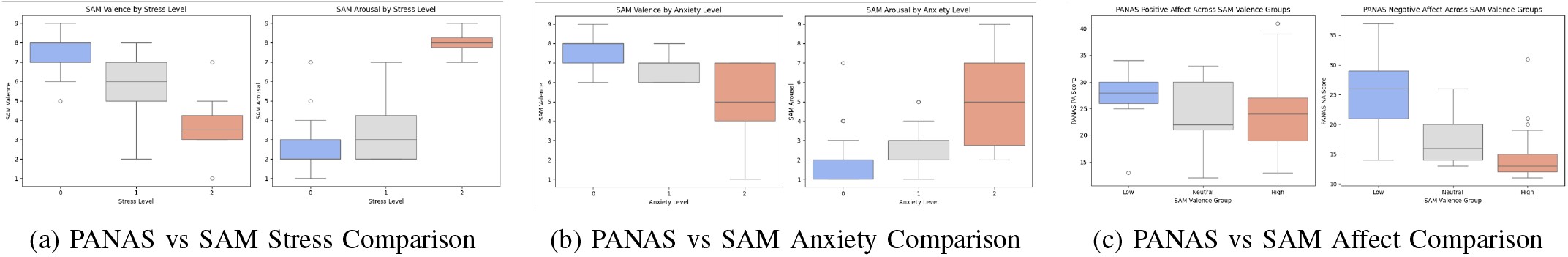
Label validation plots comparing subjective assessments from PANAS and SAM for (a) stress, (b) anxiety, and (c) overall affect.

### D. Evaluation Metrics

Classification tasks were evaluated using Accuracy, F1-Score, and AUC-ROC. Regression tasks used *R*^2^, MSE, and MAE. All metrics were computed for both training and test sets. Feature importances were computed for top-performing models.

## IV. Results and Discussion

### A. Stress Level Prediction

The static Random Forest model demonstrated limited ability to distinguish between stress levels, with AUC values of 0.55, 0.57, and 0.69 for classes 0, 1, and 2, respectively (Figure 4d). The confusion matrix showed frequent misclassification of No Stress as Low Stress (52 out of 108 class 0 samples). In contrast, the time-series LightGBM model correctly classified 84 of 88 No Stress samples and achieved AUC values of 0.94, 0.93, and 0.98 for classes 0, 1, and 2. These results confirm that incorporating recent historical physiological data enhances the model’s ability to capture subtle temporal changes in stress levels.

**Fig. 4:**
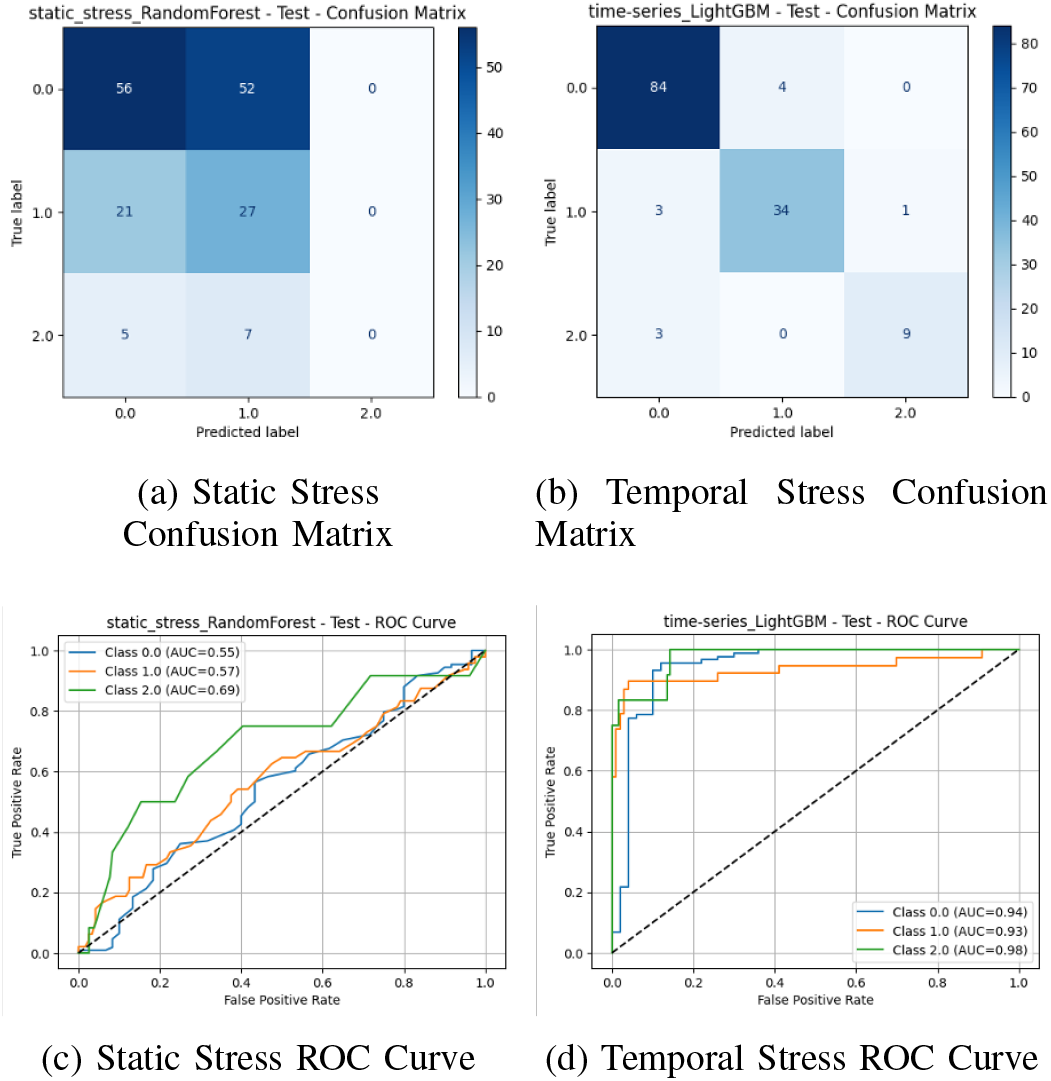
Comparative performance analysis of static and temporal stress prediction models. Subfigures (a) and (b) display the confusion matrices evaluating classification accuracy, while (c) and (d) illustrate the corresponding Receiver Operating Characteristic (ROC) curves for the static and temporal approaches, respectively.

### B. Anxiety Level Prediction

The static SVM model yielded AUC scores of 0.43, 0.47, and 0.73 for Classes 0, 1, and 2 (Figure 5). The time-series LightGBM model achieved significantly higher AUC scores of 0.93, 0.94, and 0.97, demonstrating excellent discriminative performance across all anxiety categories and confirming that temporal context substantially improves anxiety classification.

**Fig. 5:**
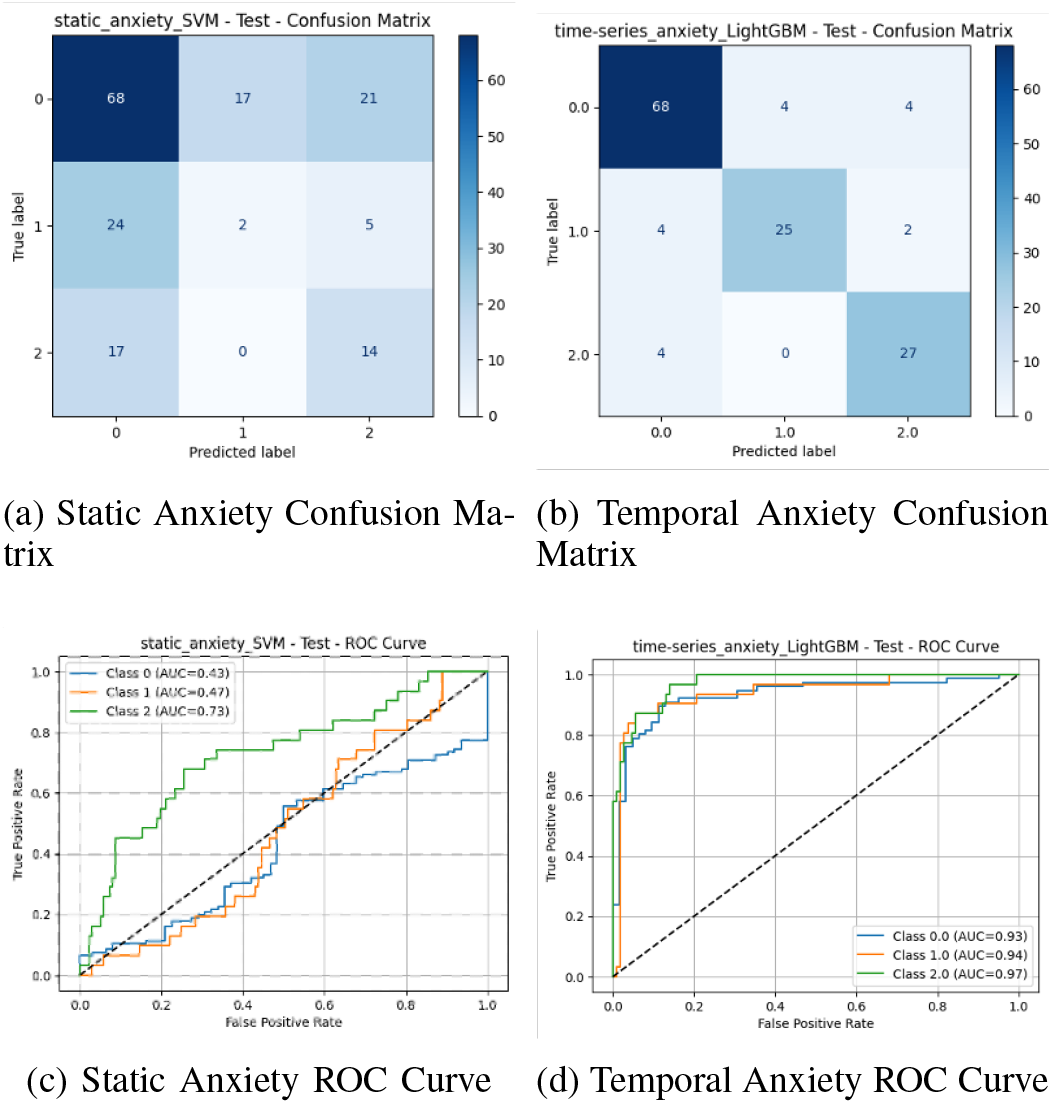
Comparative performance analysis of static and temporal anxiety prediction models. Subfigures (a) and (b) present the confusion matrices detailing classification accuracy, while (c) and (d) depict the corresponding Receiver Operating Characteristic (ROC) curves for the static and temporal approaches, respectively.

### C. Affective State Prediction

For Positive Affect, the static LightGBM model resulted in a negative *R*^2^ on the test set, indicating worse performance than a mean-value predictor (Figure 6b). The time-series Linear Regression model achieved a test *R*^2^ of 0.61, highlighting the advantage of temporal context. The model captures individual differences in affect and predictions closely follow the identity line, especially for moderate-to-high PA scores.

**Fig. 6:**
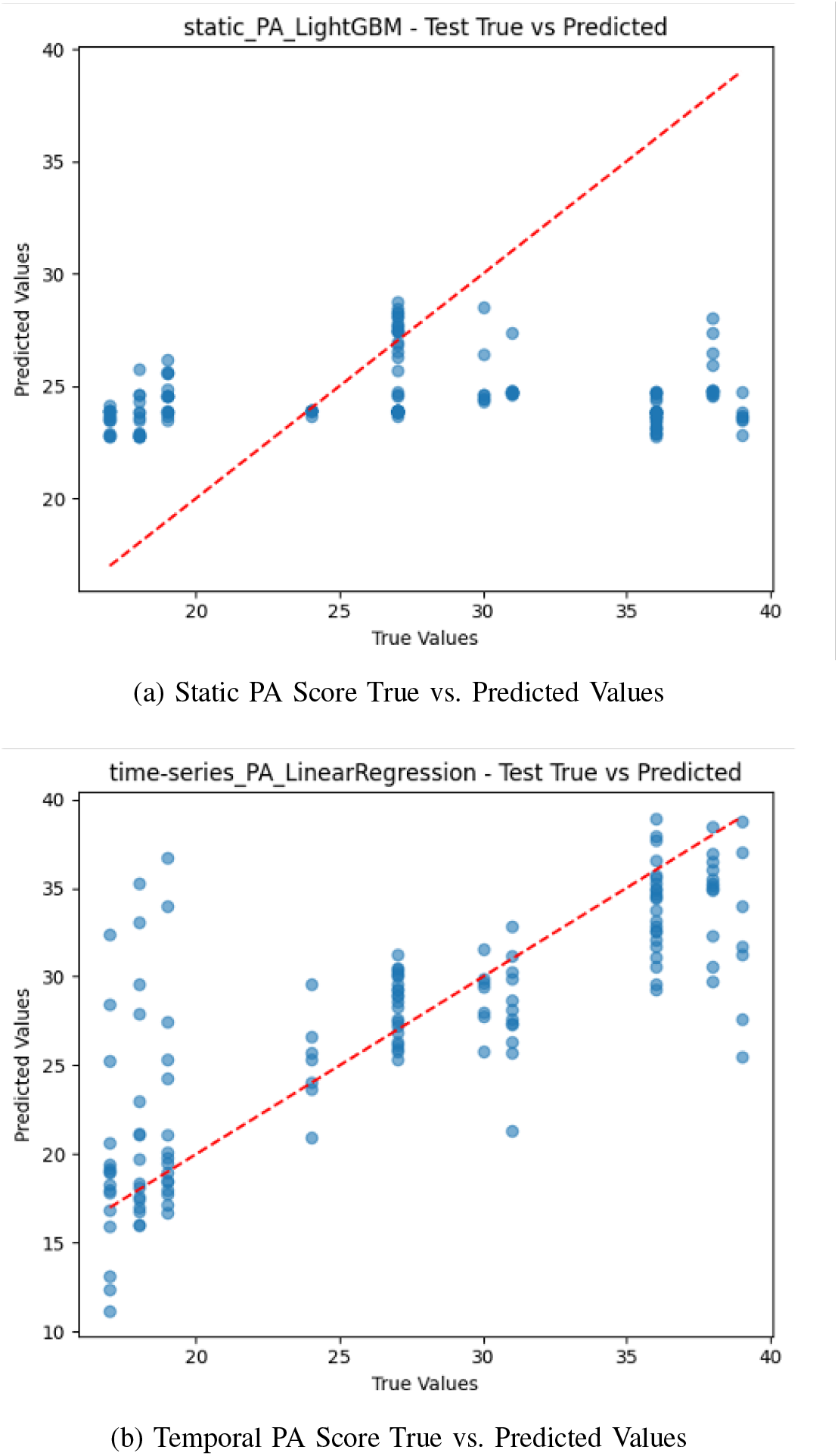
Comparative analysis of Positive Affect (PA) score predictions. The scatter plots illustrate the relationship between true and predicted values for (a) the static model and (b) the temporal model.

For Negative Affect, the static XGBoost model exhibited extreme underfitting with predictions clustering around 15 regardless of the true score, yielding a test *R*^2^ near zero (Figure 7b). The time-series SVM model performed substantially better, with predicted values more closely matching the distribution of actual NA scores, resulting in *R*^2^ of 0.68 and reduced error metrics across the board. These results show that static models fail to capture dynamic affective states, while temporal features allow for meaningful prediction of emotional variation.

**Fig. 7:**
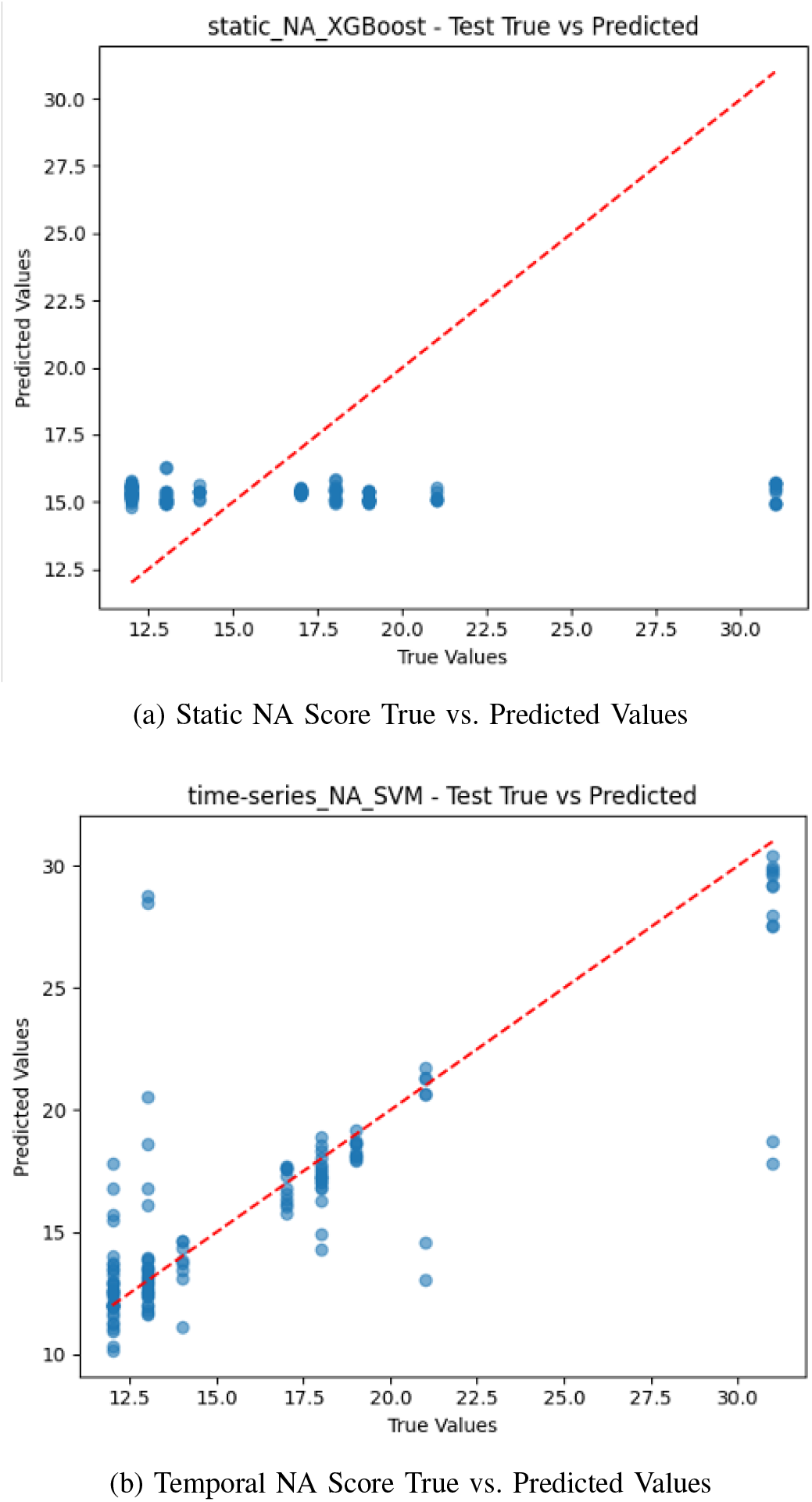
Comparative analysis of Negative Affect (NA) score predictions. The scatter plots illustrate the relationship between true and predicted values for (a) the static model and (b) the temporal model.

### D. Summary of Best Model Performance

Table II summarizes the best-performing models across all tasks. Time-series models consistently outperformed their static counterparts across all prediction tasks.

**TABLE II:** Summary of Best Model Performance.

| Task | Static Model | Time-Series Model | Static Test | TS Test |
| --- | --- | --- | --- | --- |
| Stress | RF | LightGBM | 0.49 / 0.60 | 0.92 / 0.95 |
| Anxiety | SVM | LightGBM | 0.50 / 0.54 | 0.87 / 0.95 |
| PA | LightGBM | Lin. Reg. | $R^2=-0.19$ | $R^2=0.61$ |
| NA | XGBoost | SVM | $R^2=-0.03$ | $R^2=0.68$ |

Classification metrics: Accuracy / AUC-ROC.

### E. Feature Importance Analysis

As shown in Table III, across all tasks, features derived from EDA and HRV emerged as the most influential predictors. For stress prediction, static models favored SCL Mean, HRV MedianRR, and HRV RMSSD, while time-series models favored lagged HRV-based metrics such as HRV HF t-1 and HRV LF t-2, suggesting temporal patterns in autonomic nervous system regulation are key to detecting acute stress. For anxiety, both approaches emphasized EDA and HRV complexity measures; SCL Mean, HRV pNN50, and HRV SampEN emerged as critical features in static models, while time-series models prioritized HRV HF t-1 and long-term trends like SCL Mean t-10. Temporal features such as SCR Peaks Count t-1, SCL Mean t-1, and HRV LF HF Ratio t-8 dominated time-series models for affective state prediction. Several top-ranked features included lagged values (e.g., HRV MedianRR t-9, SCL STD t-2), demonstrating that physiological trends several minutes prior hold predictive value and supporting the physiological delay between autonomic responses and subjective perception. Taken together, these findings confirm that HRV high-frequency power (HRV HF) and skin conductance mean (SCL Mean) are key indicators of emotional and stress states. The recurrent importance of lagged features in time-series models further validates the hypothesis that historical physiological context enhances prediction accuracy for real-time mental health monitoring.

**TABLE III:**
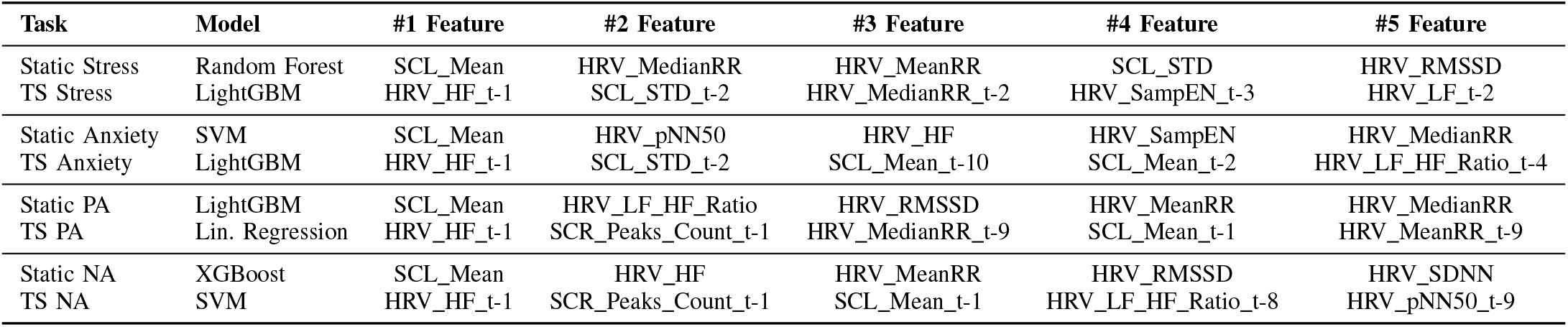
Top Five Features by Task and Model.

| Task | Model | #1 Feature | #2 Feature | #3 Feature | #4 Feature | #5 Feature |
| --- | --- | --- | --- | --- | --- | --- |
| Static Stress | Random Forest | SCL_Mean | HRV_MedianRR | HRV_MeanRR | SCL_STD | HRV_RMSSD |
| TS Stress | LightGBM | HRV_HF_t-1 | SCL_STD_t-2 | HRV_MedianRR_t-2 | HRV_SampEN_t-3 | HRV_LF_t-2 |
| Static Anxiety | SVM | SCL_Mean | HRV_pNN50 | HRV_HF | HRV_SampEN | HRV_MedianRR |
| TS Anxiety | LightGBM | HRV_HF_t-1 | SCL_STD_t-2 | SCL_Mean_t-10 | SCL_Mean_t-2 | HRV_LF_HF_Ratio_t-4 |
| Static PA | LightGBM | SCL_Mean | HRV_LF_HF_Ratio | HRV_RMSSD | HRV_MeanRR | HRV_MedianRR |
| TS PA | Lin. Regression | HRV_HF_t-1 | SCR_Peaks_Count_t-1 | HRV_MedianRR_t-9 | SCL_Mean_t-1 | HRV_MeanRR_t-9 |
| Static NA | XGBoost | SCL_Mean | HRV_HF | HRV_MeanRR | HRV_RMSSD | HRV_SDNN |
| TS NA | SVM | HRV_HF_t-1 | SCR_Peaks_Count_t-1 | SCL_Mean_t-1 | HRV_LF_HF_Ratio_t-8 | HRV_pNN50_t-9 |

### F. Limitations

Several limitations should be acknowledged. First, the WESAD dataset includes only 15 participants with a majority being male and within a narrow age range (24–35), restricting generalizability and preventing fairness analysis across demographic subgroups. Second, time-series model improvements may partially reflect overfitting to laboratory-induced transitions rather than naturalistic fluctuations. A notable unexpected outcome was the poor performance of PA regression; upon validation with SAM valence ratings, PANAS PA scores did not reliably correspond with high-valence self-reports, suggesting PANAS may not effectively capture transient emotional states in controlled experimental settings with short timeframes.

## V. Conclusions and Future Work

This paper demonstrates the feasibility of predicting minute-by-minute psychological states using wearable sensor data and time-series machine learning models. By leveraging physiological signals such as EDA and HRV, the proposed models successfully forecasted self-reported stress, anxiety, and affective states, with time-series models consistently outperforming static counterparts. Stress and anxiety classification models achieved test accuracies above 85%, and time-series regression models for negative affect achieved an *R*^2^ of 0.68, highlighting the value of temporal context in modeling psychological states. The results reinforce the importance of temporal context of physiological data for accurate emotional state prediction. HRV-derived features; particularly HF and RMSSD, along with EDA variability, emerged as consistent top predictors across tasks, aligning with existing literature on stress and autonomic nervous system responses. However, the poor alignment between PANAS PA scores and momentary valence, as indicated by SAM validation, underscores the need for improved affect labeling strategies. This outcome highlights a broader issue in affective computing: the limitations of using coarse or trait-like self-report measures for state-level prediction. Future directions include: expanding validation to real-world settings to evaluate generalization, enhancing affect measurement through real-time self-report methods or continuous valence-arousal mapping, assessing model fairness across demographic subgroups with larger and more diverse datasets, and exploring deep learning architectures such as LSTM or transformers for capturing nonlinear temporal dependencies and improving affective state prediction. Overall, this study provides a promising foundation for wearable-based mental health monitoring and real-time intervention systems.

## Data Availability

All data produced in the present study are available upon reasonable request to the authors

## References

[1] Substance Abuse and Mental Health Services Administration, “2022 national survey on drug use and health,” Tech. Rep., 2023.

[2] E. P. Terlizzi and B. Zablotsky, “Symptoms of anxiety and depression among adults: United states, 2019 and 2022,” CDC, Tech. Rep., 2024.

[3] Y.-Z. Liu, Y.-X. Wang, and C.-L. Jiang, “Inflammation: The common pathway of stress-related diseases,” Front. Hum. Neurosci., vol. 11, p. 316, Jun. 2017.

[4] M. R. Salleh, “Life event, stress and illness,” Malays. J. Med. Sci., vol. 15, no. 4, pp. 9–18, Oct. 2008.

[5] A. Holmes, “G2B reviews: stress at the intersection of anxiety, addiction and eating disorders,” Genes Brain Behav., vol. 14, no. 1, pp. 1–3, Jan. 2015.

[6] L. A. Clark, B. Cuthbert, R. Lewis-Fernández, W. E. Narrow, and M. Reed, “Three approaches to understanding and classifying mental disorder: ICD-11, DSM-5, and the national institute of mental health’s research domain criteria (RDoC),” Psychol Sci Public Interest, vol. 18, no. 2, pp. 72–145, Sep. 2017.

[7] M. Benchekroun, P. E. Velmovitsky, D. Istrate, V. Zalc, P. P. Morita, and D. Lenne, “Cross dataset analysis for generalizability of HRV-based stress detection models,” Sensors (Basel), vol. 23, no. 4, p. 1807, Feb. 2023.

[8] X. Yu, J. Lu, W. Liu, Z. Cheng, and G. Xiao, “Exploring physiological stress response evoked by passive translational acceleration in healthy adults: a pilot study utilizing electrodermal activity and heart rate variability measurements,” Sci. Rep., vol. 14, no. 1, p. 11349, May 2024.

[9] E. Lazarou and T. P. Exarchos, “Predicting stress levels using physiological data: Real-time stress prediction models utilizing wearable devices,” AIMS Neurosci, vol. 11, no. 2, pp. 76–102, Apr. 2024.

[10] M. Abd Al-Alim, R. Mubarak, N. M. Salem, and I. Sadek, “A machine-learning approach for stress detection using wearable sensors in free-living environments,” Computers in Biology and Medicine, vol. 179, p. 108918, 2024. [Online]. Available: https://www.sciencedirect.com/science/article/pii/S0010482524010035

[11] P. Schmidt, A. Reiss, R. Duerichen, C. Marberger, and K. Van Laerhoven, “Introducing wesad, a multimodal dataset for wearable stress and affect detection,” in Proceedings of the 20th ACM International Conference on Multimodal Interaction, ser. ICMI ‘18. New York, NY, USA: Association for Computing Machinery, 2018, p. 400–408. [Online]. Available: 10.1145/3242969.3242985

[12] D. Makowski, T. Pham, Z. J. Lau, J. C. Brammer, F. Lespinasse, H. Pham, C. Schölzel, and S. H. A. Chen, “NeuroKit2: A python toolbox for neurophysiological signal processing,” Behavior Research Methods, vol. 53, no. 4, pp. 1689–1696, feb 2021. [Online]. Available: 10.3758%2Fs13428-020-01516-y

[13] V. Rossi and G. Pourtois, “Transient state-dependent fluctuations in anxiety measured using STAI, POMS, PANAS or VAS: a comparative review,” Anxiety Stress Coping, vol. 25, no. 6, pp. 603–645, Aug. 2011.

[14] O. Kayikcioglu, S. Bilgin, G. Seymenoglu, and A. Deveci, “State and trait anxiety scores of patients receiving intravitreal injections,” Biomed. Hub, vol. 2, no. 2, pp. 1–5, May 2017.

[15] J. R. Crawford and J. D. Henry, “The positive and negative affect schedule (PANAS): construct validity, measurement properties and normative data in a large non-clinical sample,” Br. J. Clin. Psychol., vol. 43, no. Pt 3, pp. 245–265, Sep. 2004.

